# Re-evaluating the Cross-Sectional Prevalence of Severe Age-Related Hearing Loss Using Extreme Value Statistics

**DOI:** 10.64898/2026.06.15.26355680

**Authors:** Stefan Bleeck

**Affiliations:** Institute of Sound and Vibration Research (ISVR), University of Southampton

## Abstract

Standard demographic models of age-related hearing loss (presbycusis) predominantly utilize symmetric functions, such as log-normal distributions for age-binned thresholds and 4-parameter logistic curves for prevalence estimates. While these models capture early-to-moderate degradation effectively, they structurally struggle to characterize the heavy tails associated with severe clinical impairment. In this study, we present a statistical critique using a secondary analysis of the historical Medical Research Council (MRC) National Study of Hearing (1980–1986) dataset. By applying Generalized Extreme Value (GEV) distribution theory, we demonstrate that as severity increases, the underlying statistical geometry of hearing loss shifts. The asymmetric, heavy-tailed GEV distribution provides a parsimonious description of severe impairment, requiring fewer parameters than standard symmetric models. However, we explicitly acknowledge that utilizing static population data to infer progression introduces an ecological fallacy. Furthermore, the dataset’s historical nature embeds unquantified generational cohort effects. We conclude that while extreme value statistics offer a compelling mathematical framework for modeling the variance of severe presbycusis, true longitudinal datasets are required to isolate physiological degradation from historical cohort variance.

## 1 Introduction: Competing Statistical Models of Auditory Degradation

The prevalence of age-related hearing loss (ARHL), or presbycusis, is classically quantified by assessing audiometric thresholds across aging populations. A prominent phenomenological approach has been to describe cross-sectional threshold distributions using log-normal functions and to model the prevalence across age groups using a 4-parameter logistic curve.

While this approach yields visually satisfactory fits for mild impairments, it relies on strict mathematical assumptions. The 4-parameter logistic model assumes a symmetric, continuous probability density, where the acceleration of prevalence in middle age mirrors its deceleration in advanced age.

However, standard Normal and Log-Normal distributions arise from the Central Limit Theorem, which models outcomes as the sum of independent, uniformly distributed events. We propose an alternative statistical framework for the most severely impaired populations: that the manifestation of severe hearing loss is better modeled by extreme value statistics.

Under this hypothesis, the underlying probability density function of severe clinical impairment is governed by the Fisher-Tippett-Gnedenko theorem, and is better described by a Generalized Extreme Value Distribution (GEV). This approach utilizes a heavy-tailed, asymmetric survival geometry to capture the variance of the most affected individuals in a population without requiring artificial data shifting. While translating macroscopic population variance directly into individual biological trajectories requires extreme caution, the intuition for applying this specific statistical geometry stems from microscopic models of auditory neural activity <u>(Bleeck and Winter, 2007).</u> In that context, extreme value statistics successfully modeled neuronal spike intervals. We cite this prior work not to assert a direct physiological linkage from cellular breakdown to population distributions, but as the mathematical inspiration for hypothesizing that macroscopic clinical failures might be similarly governed by boundary-event statistics.

## 2 Methods and Analytical Framework

To evaluate whether cross-sectional ARHL prevalence is best modeled by symmetric continuous functions (Logistic/Log-Normal) or asymmetric extreme distributions (GEV/EVD), we analyzed the original cross-sectional dataset from the National Study of Hearing (NSH) <u>(Davis, 1989).</u> Data were sanitized to eliminate unphysiological artifacts. To establish a single metric of functional impairment, the PTA across 0.5, 1, 2, and 4 kHz was calculated and averaged across both ears. Participants (*N* = 2138) were grouped into 5-year age bins (spanning ages 20 to 85) to calculate the empirical prevalence of hearing loss at distinct clinical severity thresholds.

To compare the models, we utilized the Akaike Information Criterion (AIC) <u>(Akaike, 1974).</u> The AIC provides an objective measure of relative model quality by penalizing overfitting, expressed as:

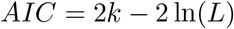

where *k* is the number of estimated parameters and *L* is the maximized value of the likelihood function. The logistic model utilizes four parameters, while the EVD model utilizes only three.

## 3 Results

The statistical analysis evaluated the cross-sectional geometry of the data across severity thresholds. First, cross-sectional fitting of the underlying PTA distribution within discrete 10-year age groups demonstrated that the 3-parameter GEV distribution captured the heavy right tail of the empirical variance more accurately than the log-normal distribution (Total AIC: 21205.3 vs 21275.6).

Second, the models were fitted to the age-binned prevalence data across severity thresholds. At the ≥ 25 dB HL clinical boundary, the continuous logistic model and the EVD model produced practically indistinguishable fits, with a Δ*AIC* of 0.21 (Logistic 66.98 vs EVD 67.19). This indicates that standard symmetric modeling remains adequate for describing the onset of mild-to-moderate impairment.

However, as the severity threshold increased, the underlying statistical geometry shifted. For severe impairment thresholds (*>* 60 dB HL), the 3-parameter EVD yielded an AIC of 66.6 against the 4-parameter logistic model’s 68.2 (Δ*AIC* = 1.6).

**Figure 1.**
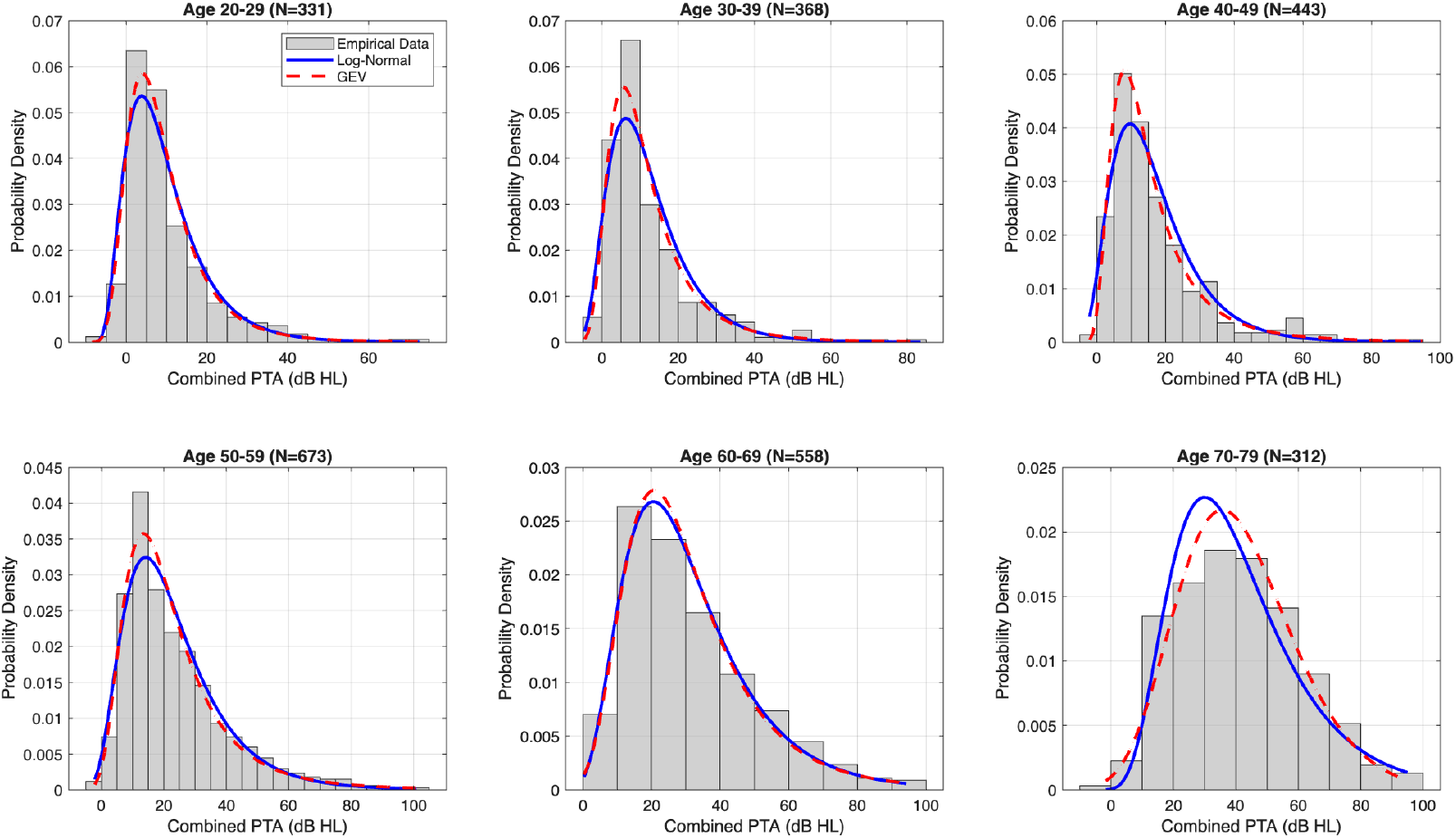
Cross-sectional probability density fits of combined PTA across 10-year age bins. The 3-parameter GEV distribution (red dashed) captures the heavy right tail of the empirical data better than the symmetric log-normal distribution (blue solid).

**Figure 2.**
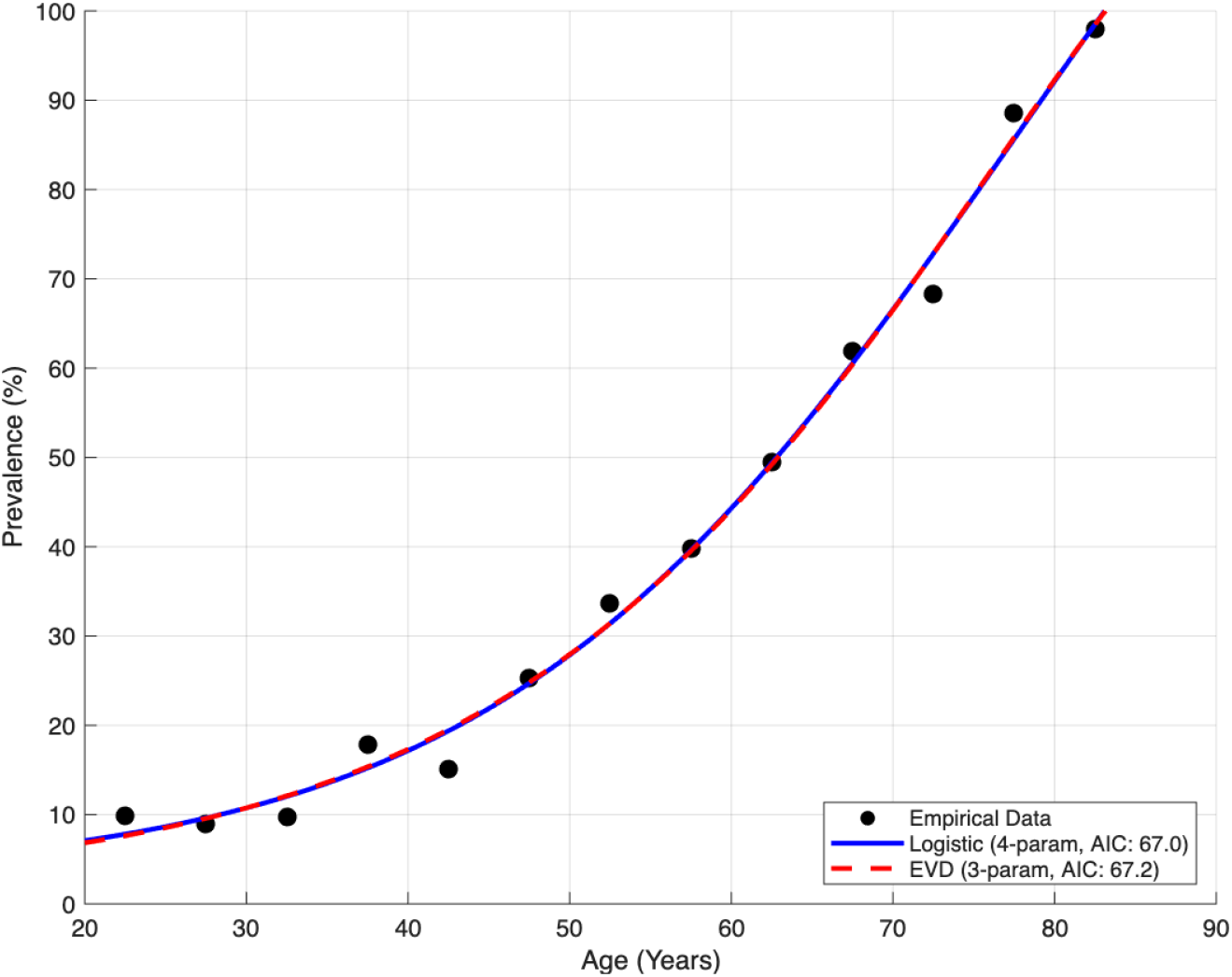
Model fit at the 25 dB HL clinical boundary. At this threshold, the continuous logistic mechanism performs similarly to the extreme-value geometry, reflecting the onset stage of impairment prior to the emergence of severe outlier tails.

**Figure 3.**
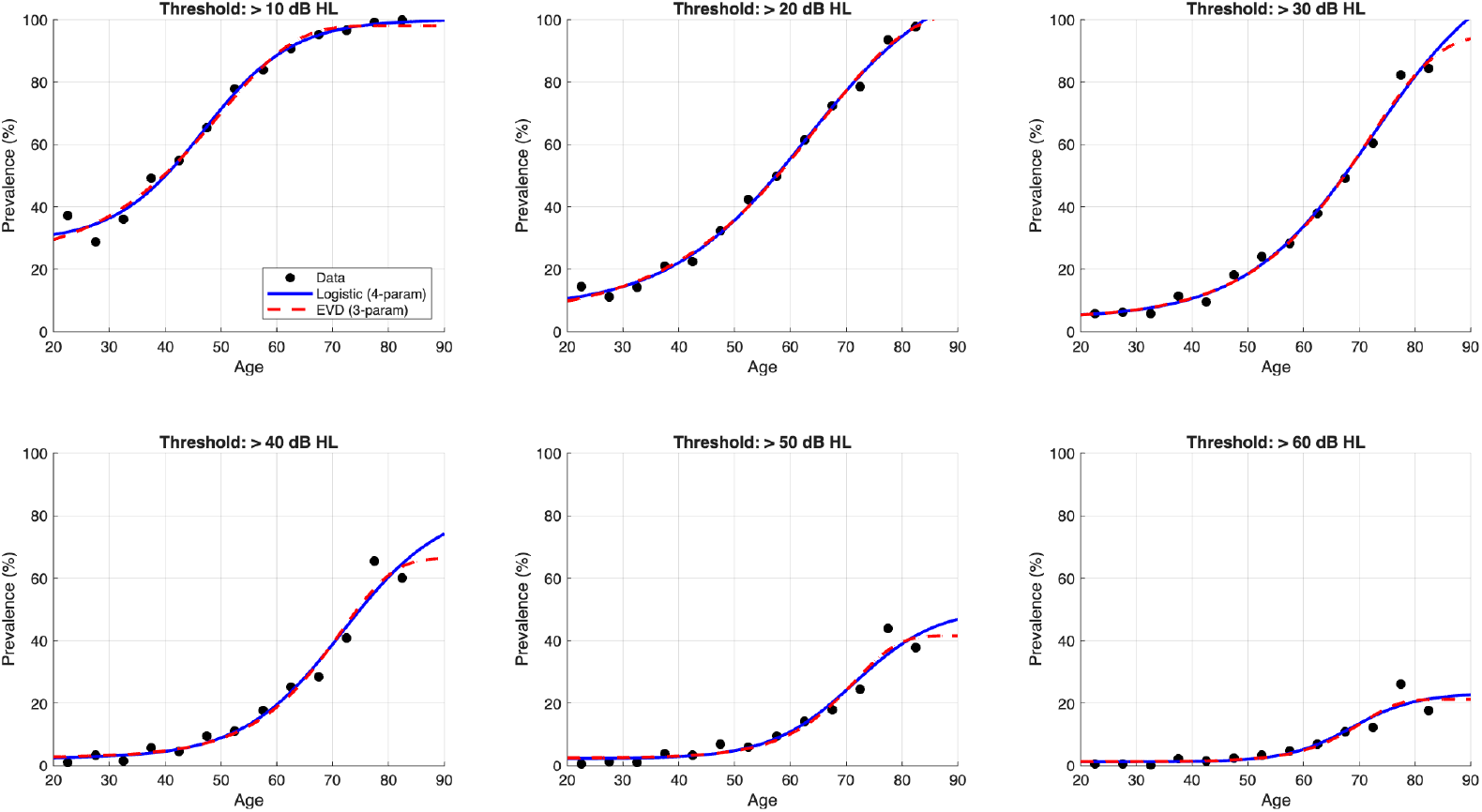
Prevalence model comparison across six severity thresholds. The symmetric logistic model (blue solid) fits lower thresholds, while the asymmetric EVD (red dashed) provides a parsimonious fit at higher, severe thresholds despite utilizing fewer parameters.

## 4 Discussion and Conclusion

The results demonstrate that standard symmetric models face mathematical limitations when describing the heavy tails associated with severe presbycusis. The observed shift in Δ*AIC* across severity thresholds suggests that the cross-sectional variance of highly impaired populations scales asymmetrically.

We explicitly acknowledge that a Δ*AIC* of 1.6 falls below the conventional threshold of 2 for definitive model superiority. However, the EVD achieves this fit utilizing one fewer parameter, representing a more parsimonious description of the data. We do not present this marginal difference as absolute proof that presbycusis is exclusively an extreme-value phenomenon. Rather, it serves as a compelling proof-of-concept demonstrating that asymmetric geometries natively capture severe threshold tails without relying on artificial data manipulation. This represents an exploratory finding that warrants future investigation, not a definitive conclusion.

Furthermore, the NSH dataset is strictly cross-sectional. Utilizing age-binned prevalence as a proxy for individual progression introduces a significant ecological fallacy. We are strictly modeling population survivability at a fixed point in time, not the longitudinal pathological trajectory of individuals.

Finally, relying on a cohort sampled between 1980 and 1986 introduces temporal validity challenges. The generational cohorts within this dataset experienced vastly different historical epochs, encompassing variations in occupational noise exposure and general healthcare standards. It cannot be assumed that the statistical geometry of presbycusis observed here is invariant to these generational cohort effects.

In conclusion, phenomenological models with high parameter counts risk overfitting structural anomalies in cross-sectional data. The Generalized Extreme Value framework provides a promising, mathematically sound approach to modeling severe impairment populations. However, isolating true physiological degradation from historical environmental variance remains impossible with cross-sectional data. True longitudinal datasets are required to validate whether this asymmetric geometry is a universal feature of auditory aging.

## Data Availability

The data analyzed in this study is a historical subset of the MRC National Study of Hearing. In accordance with institutional ethics approval (Submission ID: 113457) and university data governance guidelines, the raw dataset is restricted and kept in closed access to protect historical participant confidentiality. The aggregate mathematical parameters extracted during this analysis are fully detailed within the manuscript.

## 5 Declarations

### 5.1 Ethics Approval

This secondary data analysis was formally reviewed and approved by the Faculty Ethics Committee at the University of Southampton under Submission ID: 113457.

### 5.2 Data Availability

The data analyzed in this study is a historical subset of the MRC National Study of Hearing. In accordance with institutional ethics approval (ID: 113457) and university data governance guidelines, the raw dataset is restricted and kept in closed access to protect historical participant confidentiality.

